# The Role of Swab Tests to Decrease the Stress by COVID-19 on the Health System using AI, MLR & Statistical Analysis

**DOI:** 10.1101/2020.06.02.20120394

**Authors:** Behzad Pirouz, Hana Javadi Nejad, Galileo Violini, Behrouz Pirouz

**Affiliations:** Department of Computer Engineering, Modelling, Electronics and Systems Engineering, University of Calabria, Rende 87036, Italy; University of Calabria, Rende 87036, Italy; Centro Internacional de Física, Bogotá, Colombia; University of Calabria, Rende 87036, Italy; Department of Mechanical, Energy and Management Engineering, University of Calabria, Rende 87036, Italy

**Keywords:** Statistical analysis, Artificial Intelligence, ANN, MLR, COVID-19, Health Systems

## Abstract

The outbreak of the new Coronavirus (COVID-19) pandemic has prompted investigations on various aspects. This research aims to study the possible correlation between the numbers of swab tests and confirmed cases of infection, with special attention to the sickness level. The study is carried out with reference to the Italian case, but the result is of more general importance, in particular for countries with limited availability of ICUs (intensive care units). The statistical analysis shows correlation between the number of swab tests and those of daily positive cases, mild cases admitted to hospital, intensive care cases, recovery, and death rate, and provides a basis to carry on an AI study. The results were validated using a multivariate linear regression (MLR) approach. Our main result is the identification of a significant statistical effect of reduction of the pressure on the Health system as result of the increase of the tests. The relevance of this result is not confined to the COVID-19 outbreak, because the high demand of hospitalizations and ICU treatments due to this pandemic has an indirect effect on the possibility of guaranteeing an adequate treatment for other high-fatality disease, such as e.g. cardiological, and oncological. Our results show that swab testing may play a major role to decrease the stress on the Health system of a country. Therefore, this case study is relevant in particular for the planning of the control of the pandemic in countries with a limited capacity of admission to ICU’s units.

## 1. Introduction

The pandemic COVID-19 disease has been extensively studied considering several different aspects. Virologic studies [1] have investigated virus genoma, proving its natural origin and studying its mutations. Sociological and economical studies have considered the changes in lifestyle and the economic and sustainable development effects [2-5] correlated with non-pharmacological actions aimed to control the pandemic spread., such as lockdown [6, 7], border closure, social distancing. Other studies focused on the mechanisms and times of incubation [8], resistance and dynamics of transmission [9-11], environmental factors and conditions, like the role of UV and climate [12-15]. A large number of studies have been dedicated to statistical assessment methods and modeling of the outbreak, usually considering the number of positive cases [16-19], of active cases or of the deaths and intensive care hospitalizations [20, 21] and the health risks [22, 23].

A problem that has been object of much attention is the identification of infected people. It covers two different aspects, namely the physical identification of infected and contagious asymptomatic people, and the identification of people who have been infected and are expected to be no longer susceptible of infection. Swab testing is the usual procedure to assess experimentally the former one which otherwise could only be inferred through statistical studies [16]. In the last month in Italy, country that in the early days of the pandemic was second only to South Korea for these tests, there has been an acceleration on the use of these tests (although it should be noticed that one third of the tests was applied to people already tested at least once) and there was a significant reduction of the active cases [24].

The use of swab testing is growing in many countries as a method to fight the increasing rate of the pandemic [25, 26]. One element which is relevant for this purpose is the criterion and aim used to decide whether and to whom apply the test. Besides being a verification of recovery, this testing may be used for detection of infected people and in this case the test of random samples is opportune [17] or for the early identification of mildly infected people, which, to some extent, was actually dominant in Italy. A broad literature exists about the possibility that these tests provide a wrong answer. Gray et al. (2020) discussed the impact of this and concluded that no test would be better than a bad test [27]. In this context it must be observed that there is an important statistical difference between wrong positive and wrong negative tests [28]. Eberhardt et al. (2020) analyzed a multi-stage group testing method for testing large populations and showed that such a method would be more efficient than individual testing [29]. George and Wang (2020), analyzed the variations in testing and reporting of confirmed cases of COVID-19. Their results show the advantages of accurate scale mass and systematic tests [30]. Corman et al. (2020) determined how the tests could affect the treatment and the benefits of mild case detection [31] and to some extent our study goes in the same direction.

These studies show the importance and interplay, in COVID-19 investigation, of several variables, among which in particular swab testing. This research studies the statistical correlation between the number of swab tests and those of daily positive cases, mild cases admitted to hospital, intensive care cases, recovery, and death rate, in order to assess the role of such a systematic test for controlling the COVID-19 epidemic and reduce the pressure on the health system. The result of this research allowed us to identify certain output variables for which would be convenient to use an AI approach to study their correlation. Finally, the results of these analysis were validated using a MLR technique.

## 2. Methods

We use the available COVID-19 data in Italy and six regions, to explore the possible existence of correlations between the daily number of swabs (Swabs), and those of home isolated patients, of patients admitted to hospital but in mild condition (Mild hosp), of patients treated in the intensive care units of an hospital (Int. care hosp) and of daily deaths and daily new cases.

The approach of the study is based on the p-value, Beta coefficient, and T-test. First, in order to analyze the possible impact of daily swabs on the other variables, the datasets were divided into two periods. The statistical analysis was done for each of them and the results of the two analyses compared with each other. For each case study, in order to select the day of splitting the whole period in two periods, we first calculated the total average number of daily swabs, and then looked for the date when the number of daily swabs exceeded that average at least in three consecutive days. That date was selected for the splitting.

As next step, Machine Learning through ANN (Artificial Neural Network) has been used to investigate by Artificial Intelligence the correlations between the output variables of the statistical analysis. For Machine Learning, the variables were selected on the basis of the results of the statistical analysis, considering those that have the most significant relationship with the number of daily tests (Swab Tests). For the validation of the ANN model results, MLR (Multivariate Linear Regression) with the same variables of the ANN model has been used and a prediction equation provided. The flowchart of our analysis is presented in Figure 1.

**Figure 1.**
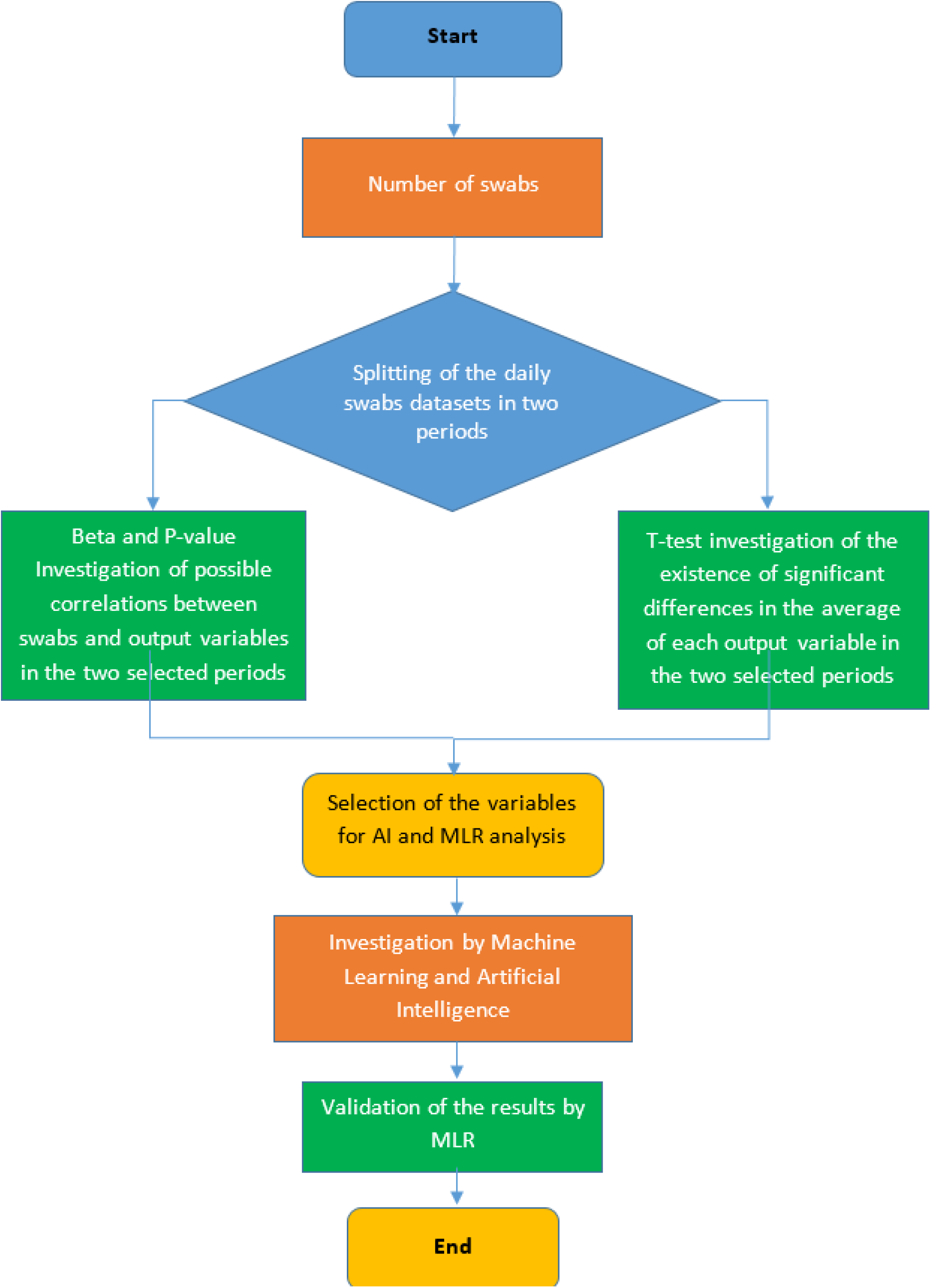
The analysis flowchart

### 2.1. Case study

In this study, we analyze the relevant datasets of six regions in Italy. Moreover, the analysis has also been carried out for the whole of Italy. The location of the six regions is shown in Figure 2 and the details of the selected case studies are presented in Appendix A.

**Figure 2.**
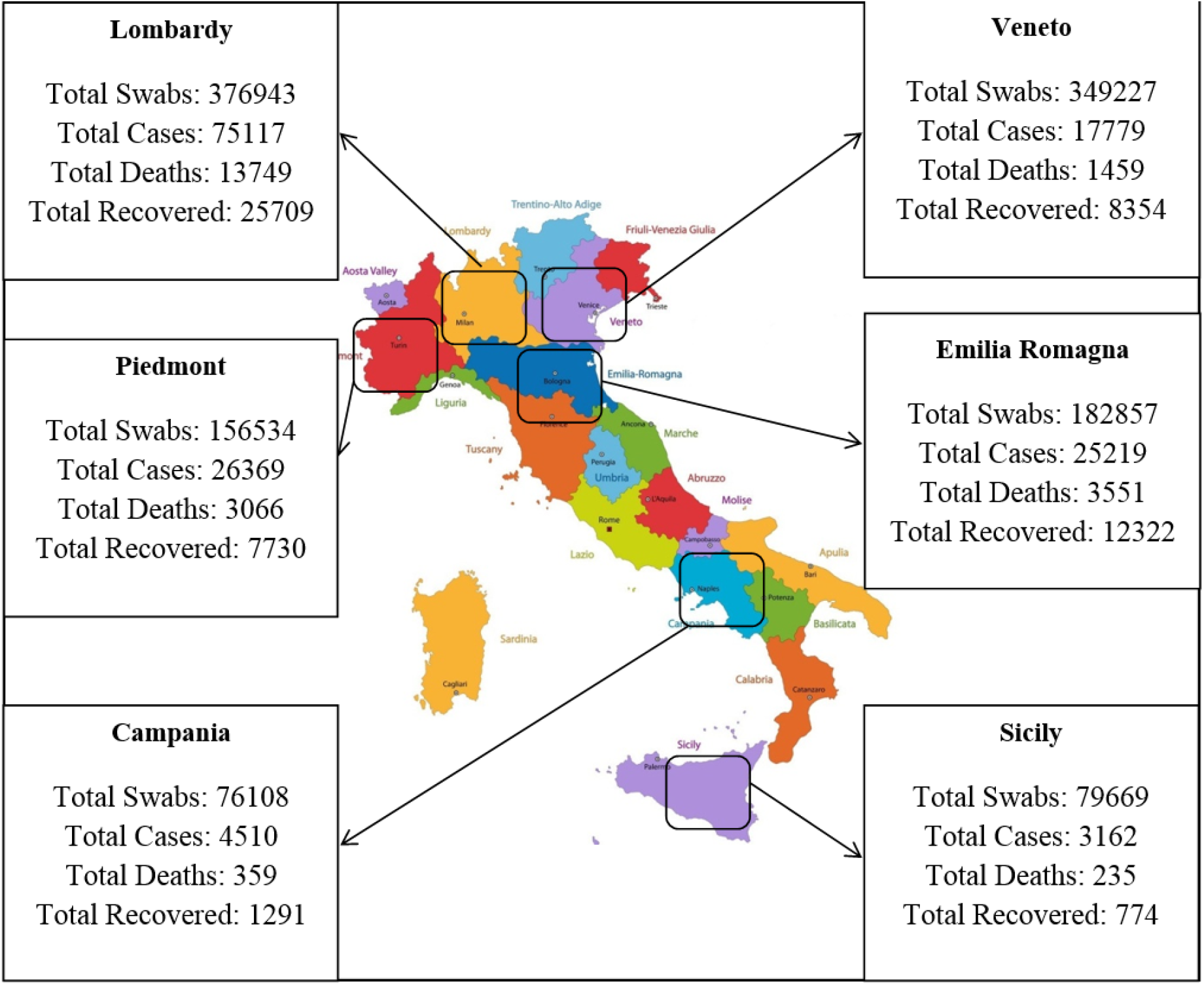
Location of the selected case studies [33-34]

### 2.2. Mathematical modeling and statistical analysis

Mathematical modeling and statistical analysis are a tool widely used in various fields, e.g. natural and human hazards [35-37] and for many phenomena, including hazards and optimization [38, 39]. In all these approaches it is essential to express the problem in form of an equation or a model [40-46]. Due to the complexity of the natural phenomena, several criteria can be used to obtain the best mathematical modeling for a given problem [47-51].

Our purpose is to study the effect of an independent variable on one or more dependent variables, and to assess the possible existence of a relationship between them. An efficient statistical technique to achieve this, is to evaluate a coefficient, the beta coefficient, that measures the degree of change in a dependent variable (outcome variable) in correspondence with a 1-unit change in the independent variable (predictor variable). Of course, the Beta coefficient value can be negative or positive. This quantitative evaluation makes sense only if the possible existence of a significant relation between the two variables has been previously assessed. This is established by evaluating another parameter, the p-value, whose value, if smaller than 0.05 (or confidence level of 95%), indicates the existence of a significant correlation between the considered variables [52]. The set-up of the SPSS Model is presented in Appendix B.

## 3. Results

### 3.1. Correlations between swabs and the other variables in whole Italy

Our first analysis refers to the global country dataset. The results of the correlations between swabs number and the output variables in the two periods are presented in Table 1. and Table 2. The daily variations of the different parameters in whole Italy are presented in Figures 3 and 4.

**Table 1.** T-test results in two periods for whole Italy

| Case study | Variables | Date | Average | Rate of change | t-test |  | Result |
| --- | --- | --- | --- | --- | --- | --- | --- |
|  |  |  |  |  | t value | P-Value |  |
| All Italy | Test No. | 1 March-31 March | 15752 | 3.12 | -11.129 | 0.000 | significant difference |
|  |  | 1 April-30 April | 49075 |  |  |  |  |
|  | Home isolation | 1 March-31 March | 15768 | 4.57 | -16.513 | 0.000 |  |
|  |  | 1 April-30 April | 72053 |  |  |  |  |
|  | Mild hosp | 1 March-31 March | 12763 | 2.00 | -6.858 | 0.000 |  |
|  |  | 1 April-30 April | 25566 |  |  |  |  |
|  | Int care hosp | 1 March-31 March | 1986 | 1.49 | -3.457 | 0.000 |  |
|  |  | 1 April-30 April | 2975 |  |  |  |  |
|  | Daily Deaths | 1 March-31 March | 400 | 1.29 | -1.901 | 0.062 | No significant difference |
|  |  | 1 April-30 April | 518 |  |  |  |  |
|  | Daily new cases | 1 March-31 March | 3376 | 0.98 | 0.131 | 0.896 |  |
|  |  | 1 April-30 April | 3322 |  |  |  |  |

**Table 2.**
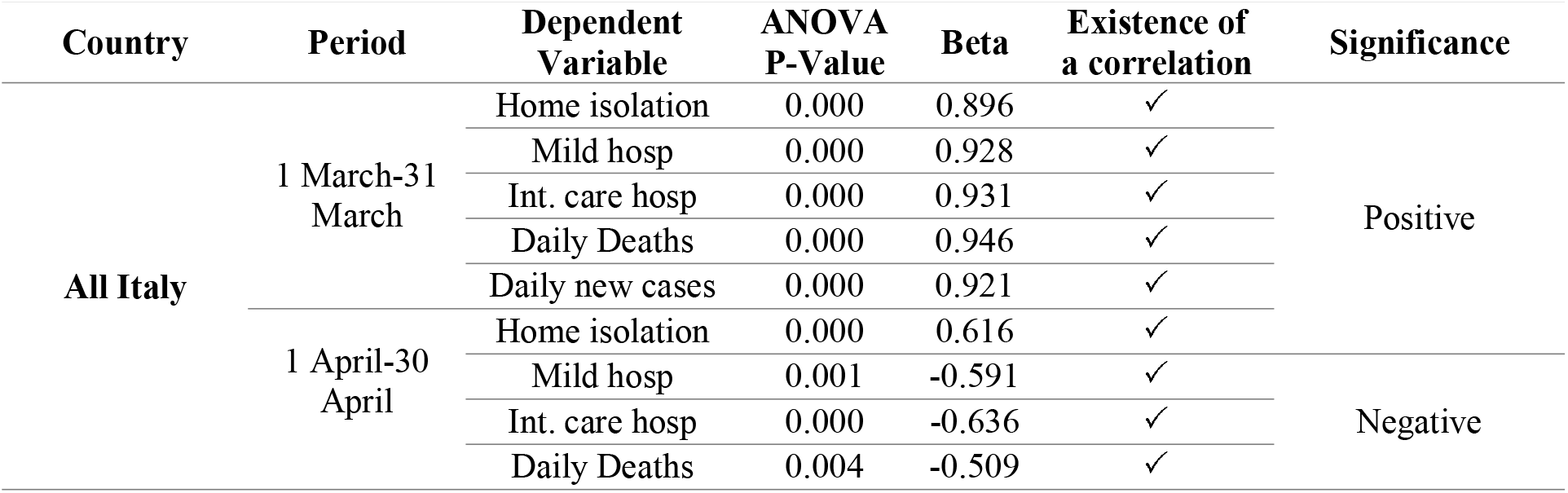
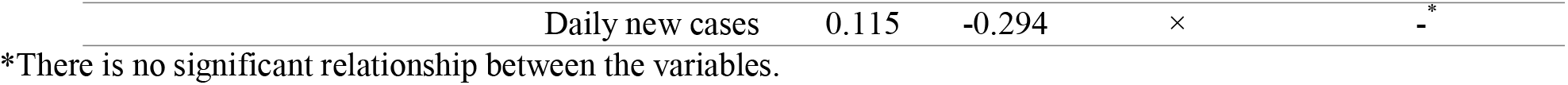
Test No. (Independent variable) Vs. each outcome variable in two time periods (All Italy)

**Figure 3.**
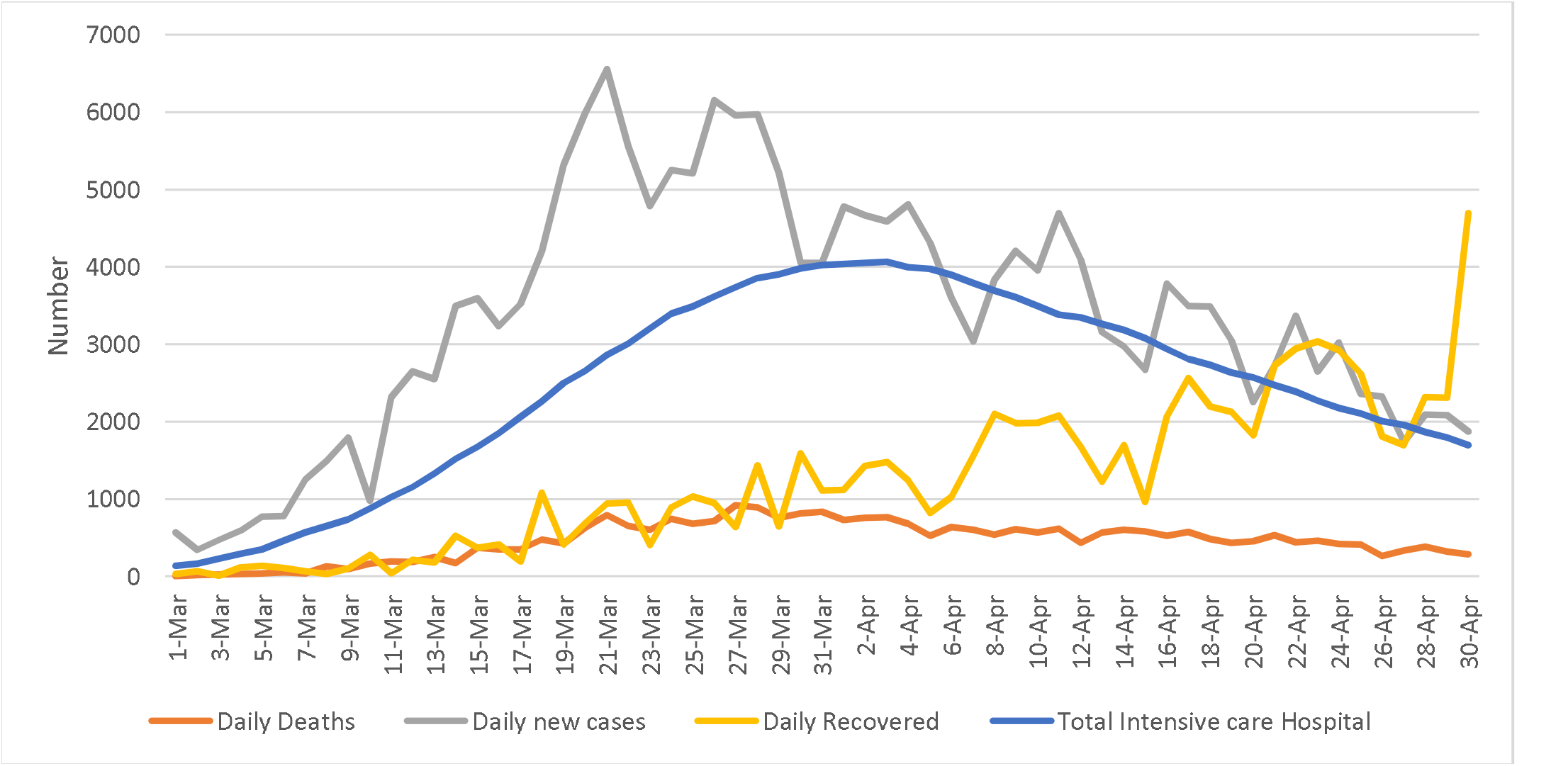
Daily variations of the parameters in whole Italy

**Figure 4.**
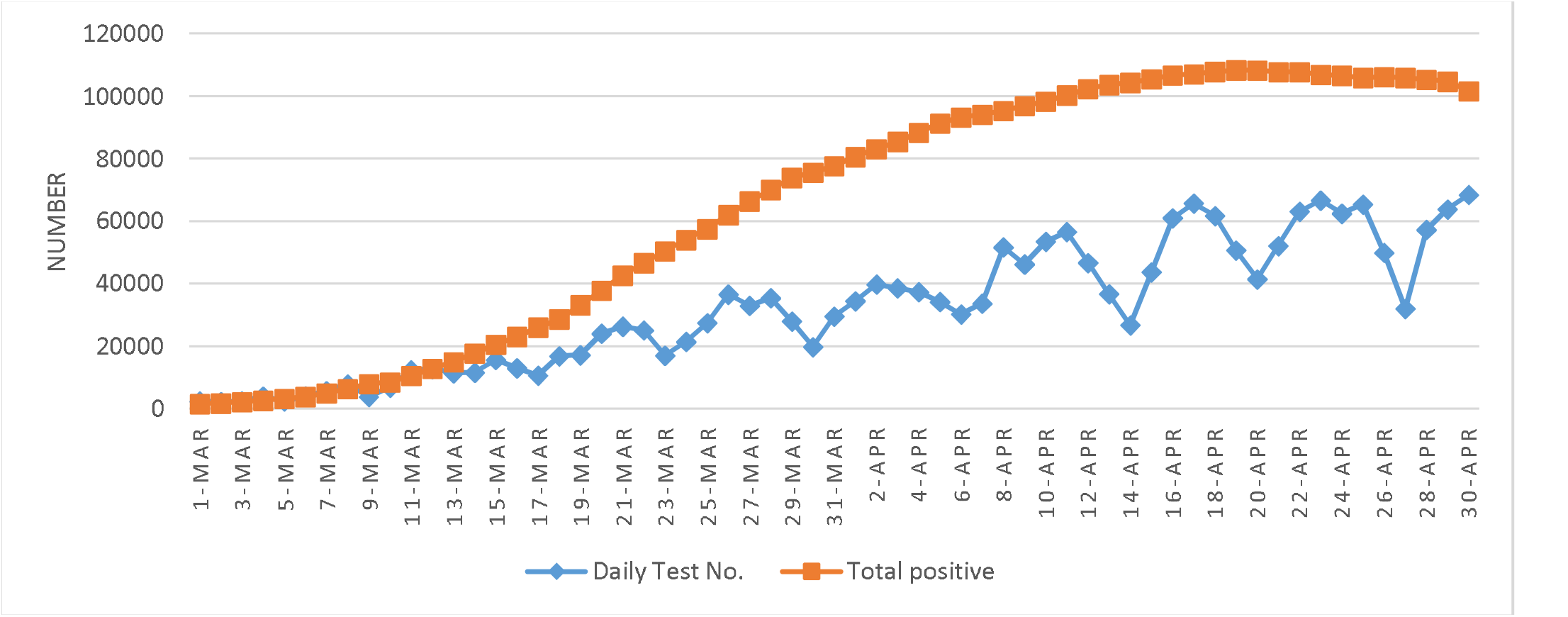
Daily tests and total positive cases in whole Italy

As shown in Table 1, in Italy the average number of the independent variable (swabs) increased significantly by 3.12 times. This is compared with the variation in the same period of the outcome variables. One sees that this increase was accompanied by a significant increase of the average number of home-isolated patients (4.57 times), Mild Hosp cases (2.00 times), and Int Care Hosp cases (1.49 times). Instead, as indicated by the corresponding p-values, the increase by a factor 1.29 in the average number of Daily Deaths, and the decrease by a factor 0.98 in the average number of Daily New Cases were not significant.

For what concerns the relationship between the daily swabs and the outcome variables, Table 2, shows that during the first period (1 March to 31 March) it was significantly positive for each of the five variables. During the second period (1 April to 30 April), the correlation with Home isolation remained significantly positive, those with Mild hosp, Int. care hosp, and Daily deaths, became significantly negative, and that with Daily new cases, although turned into negative, was not significant. The analysis at regional level presented in Appendix C.

### 3.3. Model with using ANN

As the results of Table 2. show, the number of total home isolation, total mild hospital, and intensive care in the hospital changed significantly with the number of swabs, whereas for the Daily new cases this happens only for the first period. On the other hand, increasing the number of swabs can only be related to new home isolation cases and mild hospital, without connection with that of ICU cases.

Therefore, the variables we selected for this analysis were Daily new cases, Total home isolation, and Total mild hospital, and the dependent variable was total Intensive cases in Hospital. One can suspect that there exists a correlation between them and their daily variation. In this section, we shall investigate this possible correlation using the ANN method to calculate the Intensive care number. The results of the model including train and test of the ANN algorithm are presented in Table 3. and the comparison between the prediction values and real data in Figure 5. It shows that AI is a powerful tool for the study of our problem and this is confirmed by the value of the final R^2^. Additionally, it confirms the results of the previous statistical analysis on swab-output variables correlations.

**Table 3.** The results of ANN

| Factors | Train Value | Test value |
| --- | --- | --- |
| Number of datasets | 51 | 10 |
| MSE (mean squared error) | 20613.8 | 17847.6 |
| Correct prediction | 47 | 7 |
| Wrong prediction | 4 | 3 |

For all data $R^2 = 0.986$

**Figure 5.**
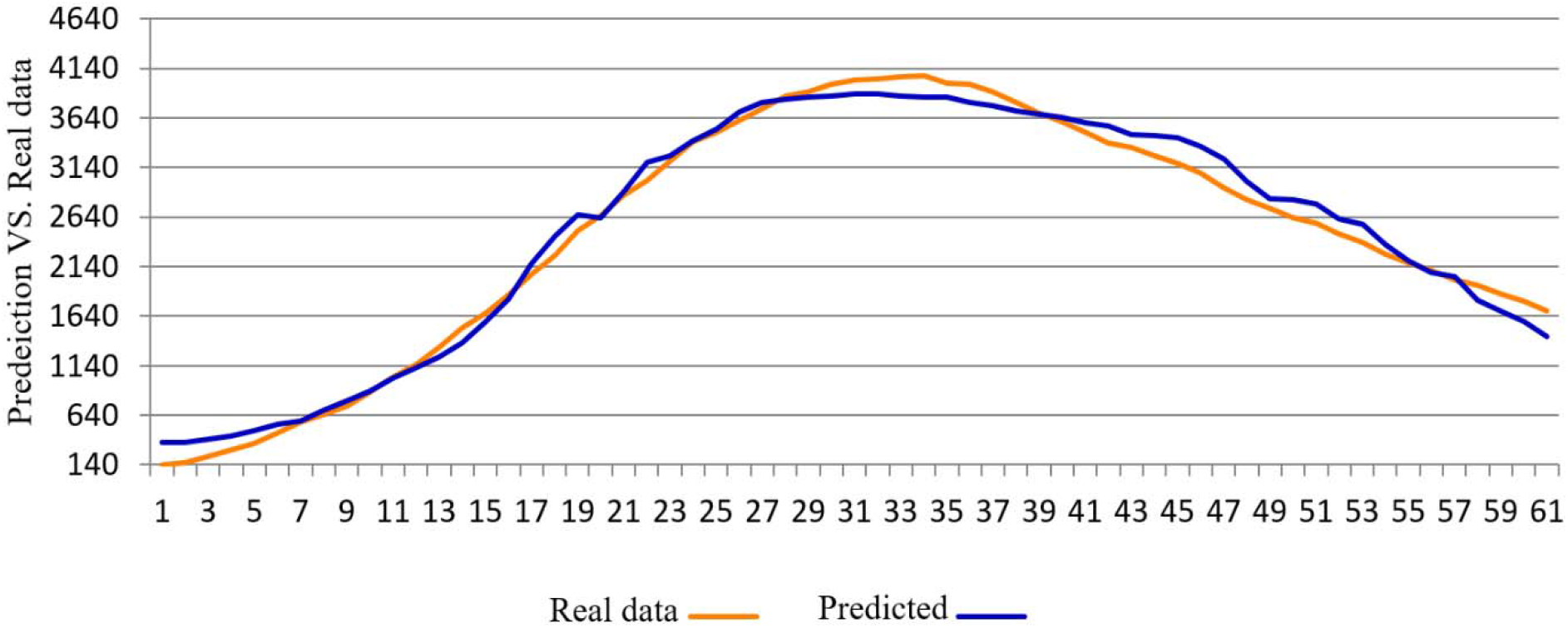
Prediction values by ANN and real data for all data

The impact of each variable on Intensive case in the hospital is shown in Figure 6. According to Figure 6, the dominance of the role of Mild hospital over home isolation for the impact on the number of intensive case in the hospital is evident.

**Figure 6.**
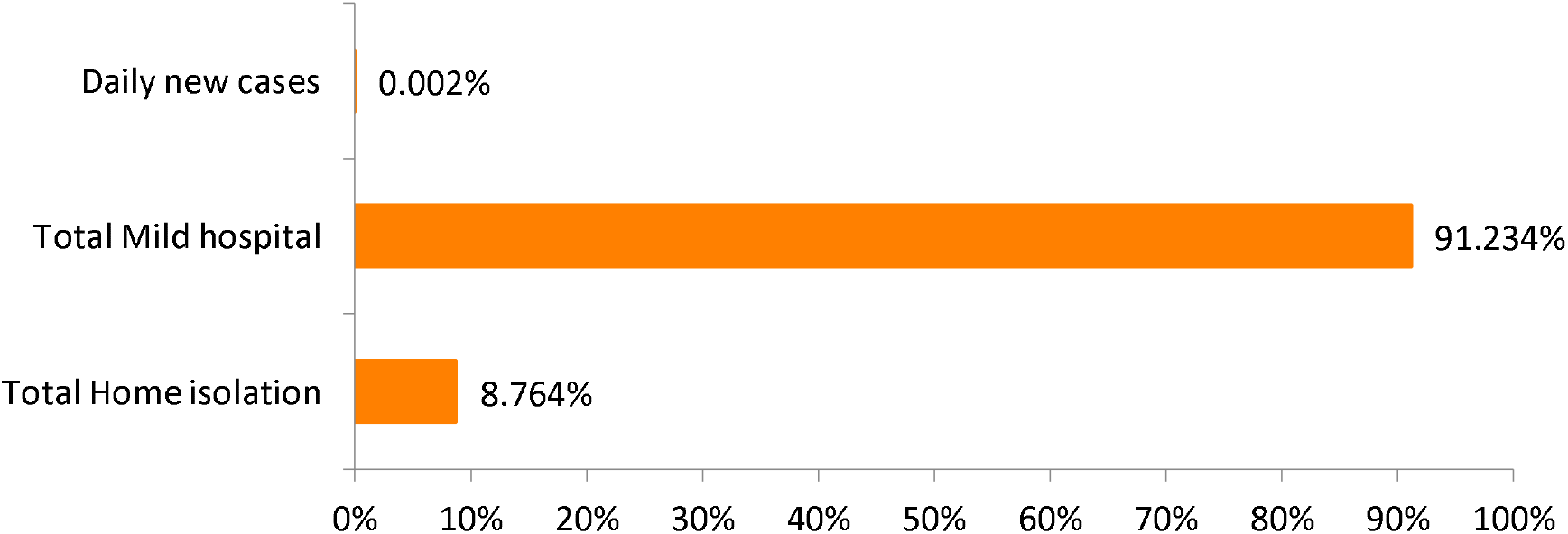
The impact of each variable on the result (Intensive case in the hospital)

## 4. Discussion

In this section we summarize the results of the previous section. The first part of our statistical analysis was based on T-test. In Italy, during the period we analyzed (March-April) there was a significant change on the number of swab tests. At country level, one can say that there was a first period (March) with a country average of about 15000 tests per day, and that this average grew more than three times in the following month. Analogous increases in the number of tests are observed at regional level, although they are probably also sensitive to the actual level of development of the pandemic in the specific region.

This preliminary analysis confirms that there was a significant change in the application of these tests. This supports the hypothesis that such a change may be reflected on the indicators that describe the level of development of the pandemic. To test this hypothesis, we carried out a statistical analysis of the possible correlation between the number of tests and that of contagious people, the recovered ones and the deaths. At national level, the hypothesis was confirmed with the exception of the daily new cases. These correlations generally are observed also at regional level, although minor deviations may occur. In particular these deviations seem to be sizeable in Emilia-Romagna and Veneto and this may find an explanation in the improvement of the situation in these two regions during the month of April [19]. This also happened with respect to the number of deaths. As it might have been expected, we found that with the increase of the number of tests in the second period the correlation between the death rate and the number of swab tests, considerably diminished with respect to the first period. Instead, significant correlations were generally found for the three variables of the outmost importance for what concerns the health system stresses related to the total number of cases, and to a less advanced level of development of the sickness. The evidence is stronger in the regions where the pandemic spread more.

Of course, this discussion must take into account the multiple reasons that lead to apply the test. This means that in any case only a fraction of those actually applied can be related to the variables we studied, but this can be estimated to be only a second order effect.

The last part of our analysis had a different purpose. Artificial Intelligence are a powerful tool to analyze a large number of data to detect possible correlations between them. We applied this technique to study one of the most critical aspects of the pandemic, the admission to ICUs. We find a very good agreement with the observed number of ICU admission. This aspect must not be underestimated, when one takes into account that the stress on ICU is probably one of the most critical factors to take into account when planning the response to the pandemic.

In the specific case of Italy, when the peak shown in Figure 5. was reached, the value of ICU hospitalizations was close to the capacity of response of the Health system, although it was planned for a couple of weeks later a 40% increase of the available ICU units [53]. We recall that Italian ICU capacity was then in the order of 120 ICUs per million population, and it must be underscored that this is a high performance, in compare with different countries as presented in Appendix D.

A validation of our results on the role of swabs is provided by the determination of a prediction equation obtained using MLR. We already pointed out the usefulness of the swabs to identify infected people without symptom who then would be home isolated. The variables for our MLR analysis are the total number of Home isolation (x_1_), that of mild condition admissions to hospital (x_2_) and that of Daily new cases (x_3_). These variables are used as independent variables to determine as dependent variable, y, the total number of patients admitted in the ICUs of a hospital.

The prediction equation we obtain is

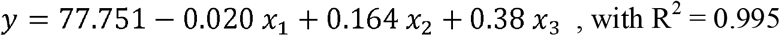

Table 4. provides the details of the analysis and the values of the beta coefficients show that the highest impact variable is x_2_ followed by the variable x_1_. We notice that the increase in the number of home isolation cases that we found to be correlated to that of swab tests is associated to a decrease of the total number of patients admitted to hospital intensive care units.

**Table 4.** The result of MLR analysis

| Variables | P-Value | Beta | Coefficients |
| --- | --- | --- | --- |
| Constant | 0.011 | - | 77.751 |
| Total Home isolation ($x_1$) | 0.000 | -0.514 | -0.020 |
| Total Mild Hospital ($x_2$) | 0.000 | 1.311 | 0.164 |
| Daily new cases ($x_3$) | 0.009 | 0.050 | 0.38 |

## 5. Conclusions

We studied the statistic correlation between the number of applied swab tests and the most important indicators of the level of development of the COVID-19 pandemic, in Italy and in six Italian regions, four chosen on the basis of the diffusion of the pandemic and the remaining two on the basis of the size of their population. This analysis was accompanied by one using AI techniques and validated through an MLR. Our results show that swab testing may play a major role to decrease the stress on the Health system of a country. Therefore, this case study is relevant in particular for the planning of the control of the pandemic in countries whose capacity of admission to ICU’s units is limited.

## Data Availability

All used data is available in this site

https://lab.gedidigital.it/gedi-visual/2020/coronavirus-i-contagi-in-italia/

## Acknowledgments

We are grateful to Luigi Grasso for an interesting discussion that stimulated this research, and to Professor Manlio Gaudioso and Dr. Agostino Gnasso for their critical reading of the manuscript.

## Appendix A

### The details of the selected case studies

Our analysis covers the period from beginning of March to the end of April. Four out of the six regions we analyzed, namely Lombardy, Piedmont, Emilia-Romagna and Veneto, are those where the outbreak has been stronger. All of them are located in Northern Italy. The remaining two, Campania and Sicily, belong to the Southern part of Italy, and have been selected because the size of their population is similar to that of three of the four Northern regions.

**Table A1.** The selected case studies in Italy [54, 55]

| Region/Country | Capital | Population | Available dataset |
| --- | --- | --- | --- |
| Lombardy | Milan | 10,040,000 | 1 March to 30 April |
| Veneto | Venice | 4,906,000 | 1 March to 30 April |
| Piedmont | Turin | 4,356,000 | 6 March to 30 April |
| Emilia-Romagna | Bologna | 4,459,000 | 6 March to 30 April |
| Campania | Naples | 5,827,000 | 7 March to 30 April |
| Sicily | Palermo | 5,000,000 | 8 March to 30 April |
| Italy | Rome | 60,317,116 | 1 March to 30 April |

## Appendix B

### The set-up of the SPSS Model

To carry on our statistical analyses, SPSS model has been used to study the following variables:

- Number of daily tests (Swabs test);
- Number of patients isolated at home (Home isolation);
- Number of patients admitted to the hospital in mild condition (Mild hosp);
- Number of patients admitted to the intensive care section of hospital (Int. care hosp);
- Number of Daily Deaths;

The analysis method proceeded as discussed in the previous section.

- Use of the Independent-Samples T-Test: The Independent-Samples T-Test is used to test the existence of significant differences (with a confidence level of 95%), between the averages of two groups of data. We recall that our dataset was splitted in two subsets on the basis of the comparison of the daily number of tests with its average over the full period. The T-test was applied both to the independent variable (Swabs) and to the dependent variables (Home isolation, Mild hosp, Int. care hosp, daily deaths, and daily new cases) in order to determine whether the comparison between the two periods of the corresponding data presents a significant difference.
- Evaluation of the p-value: If the result of p-value coming from the comparison of the independent variable (Swabs) and each dependent variable (Home isolation, Mild hosp, Int. care hosp,) is less than 0.05, then the relationship between the two variables is considered to be significant and we proceed to the calculation of the Beta coefficient. If instead the p-value is larger than 0.05, the significance of the relationship is excluded.
- Evaluation of the Beta coefficient: A positive (negative) value of this coefficient means that for every 1-unit increase (decrease) in the predictor variable (swabs number in our case), the outcome variable increases (decreases) by an amount equal to the value of the beta coefficient.

## Appendix C

### The correlations between swabs and other variables at regional level

The global analysis at country level makes interesting to check whether the global trends we found are shared also at local level in the six regions we are considering. We recall that together these six regions account for 57.3% of the country population.

We present below the results of the regional analysis of the correlations between the number of swabs and those of the outcome variables, in two different periods determined through the procedure described in Section 2. Tables C1 and C2 refer to Lombardy, Veneto and Piedmont for which data are available from March 1, and Tables C3 and C4 to Emilia-Romagna, Campania and Sicily, for which the first day for which data are available was different.

**Table C1.**
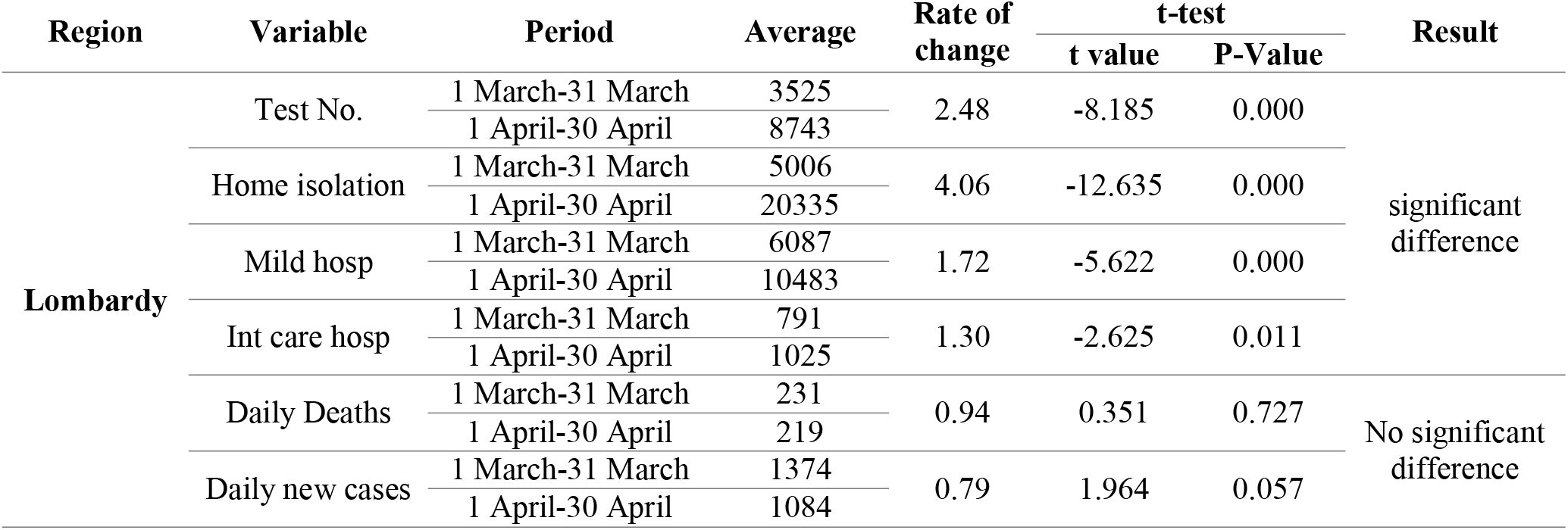
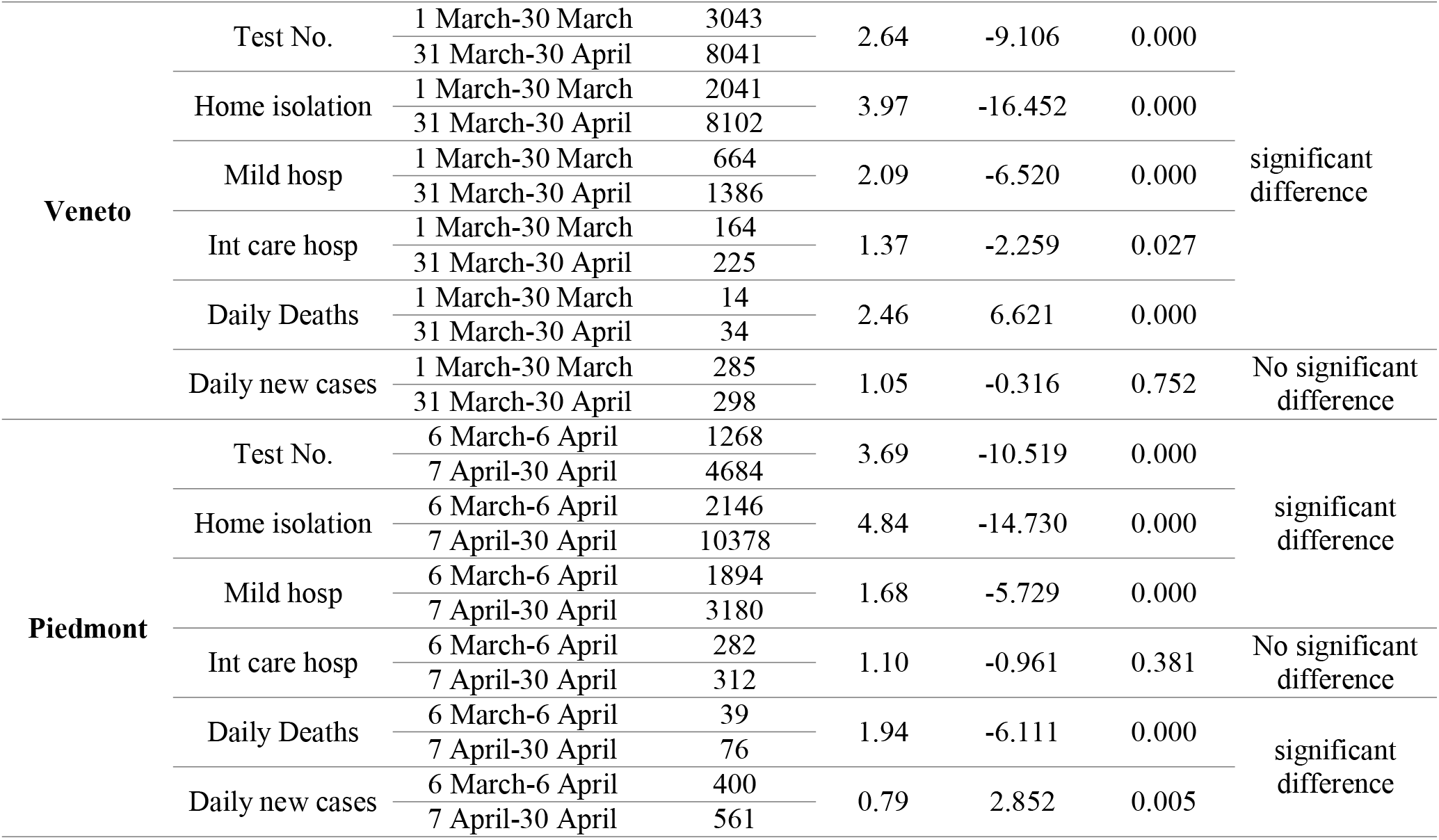
T-test results in two time periods (Lombardy, Veneto, Piedmont)

To summarize the T-test results of Table C1, we can say that:

- **In the three regions, there has been a remarkable increase in the average number of daily tests. In Lombardy by a factor 2.48, in Veneto by a factor 2.64, and in Piedmont by a factor 3.69**

Against this increase:

- The average number of home isolated and Mild Hosp increased significantly in all of them.
- The average number of Int Care Hosp also increased everywhere, although in Piedmont no significantly.
- Daily New Cases exhibit a general decrease that is not significant
- Daily Deaths decreased (not significantly) in Lombardy and significantly increased in Piedmont and Veneto.

**Table C2.**
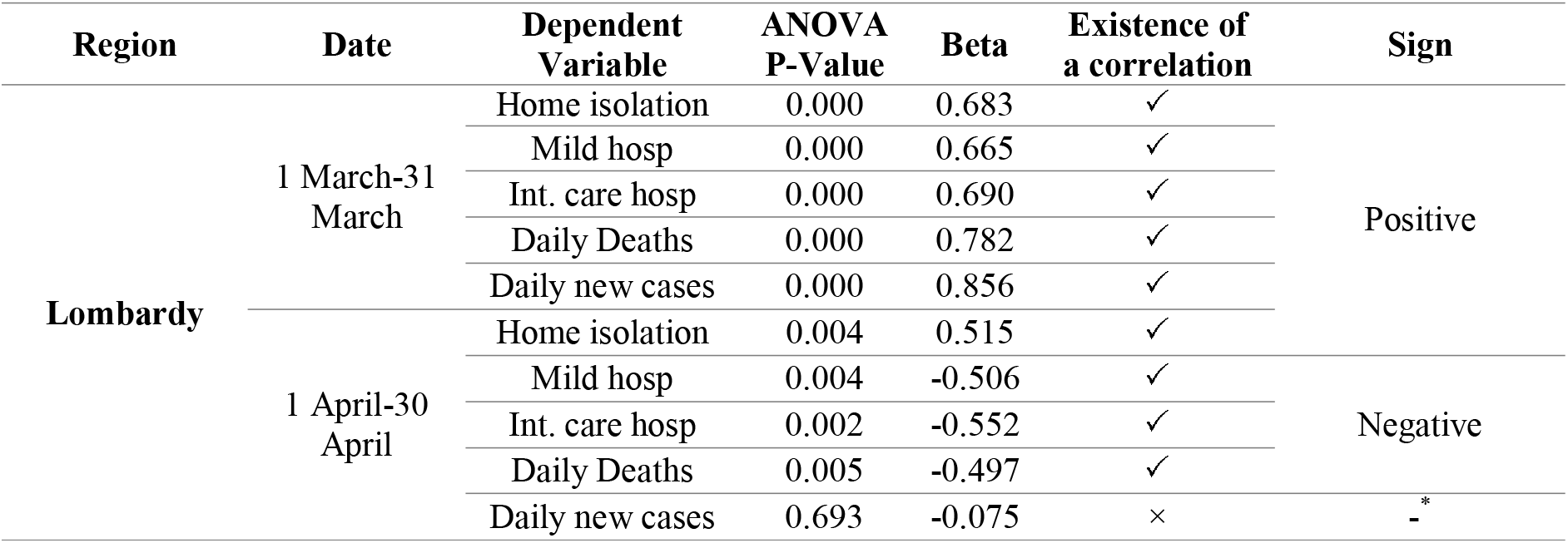
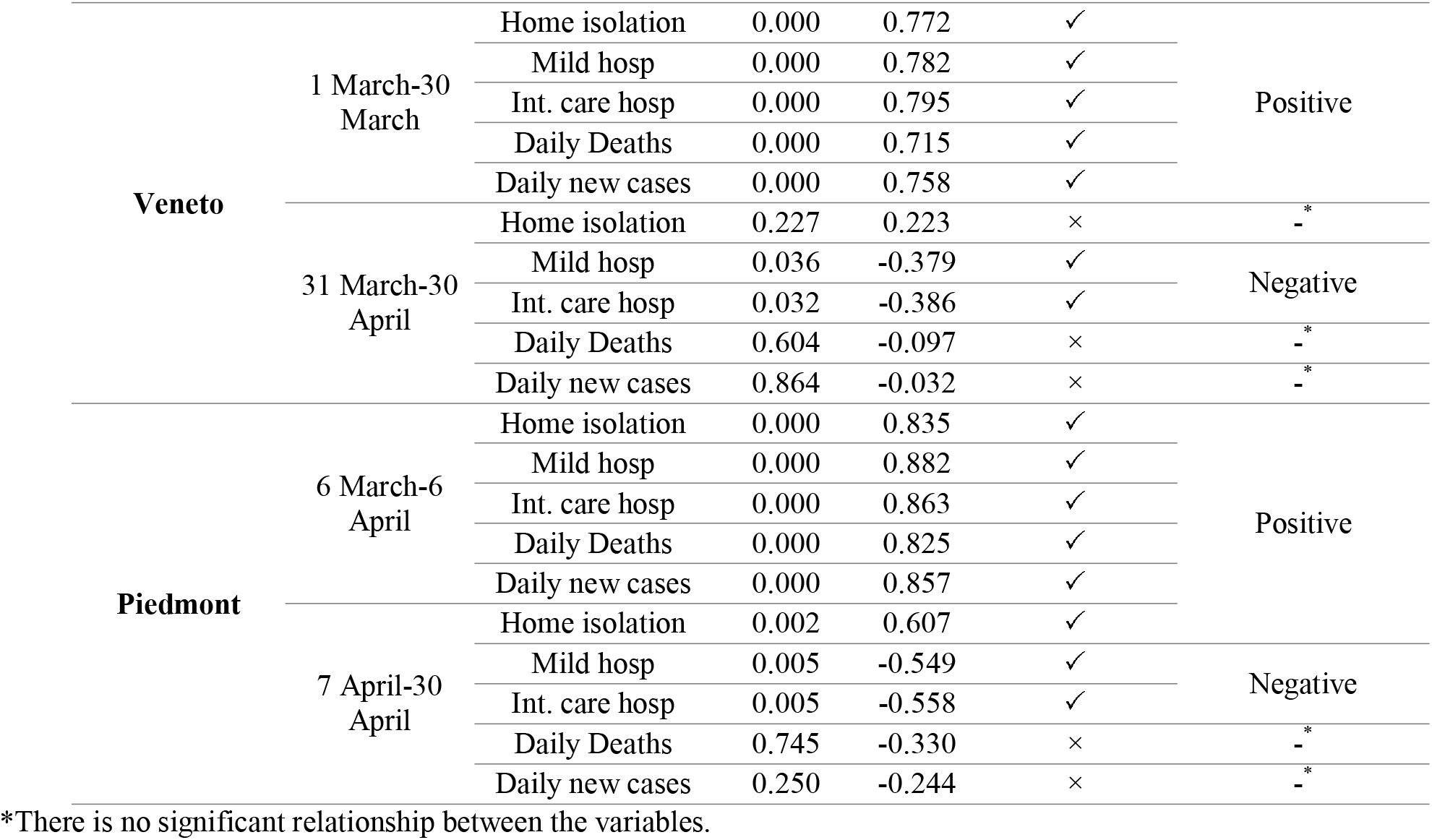
Test No. (Independent variable) Vs. each outcome variable in two time periods (Lombardy, Veneto, Piedmont)

The correlation analysis of Table C2, can be summarized as follows:

- In the first period, in each of the three regions the global Italian result was confirmed, and one finds a significantly positive correlation between the number of swabs and that of each outcome variable.
- In the second period, whereas for Home isolation there is a positive correlation, significant only in Lombardy, all the other correlations turn to negative, generally in a significant way, the only exceptions being Daily new cases in all of the three regions and Daily deaths in Veneto and Piedmont.

**Table C3.**
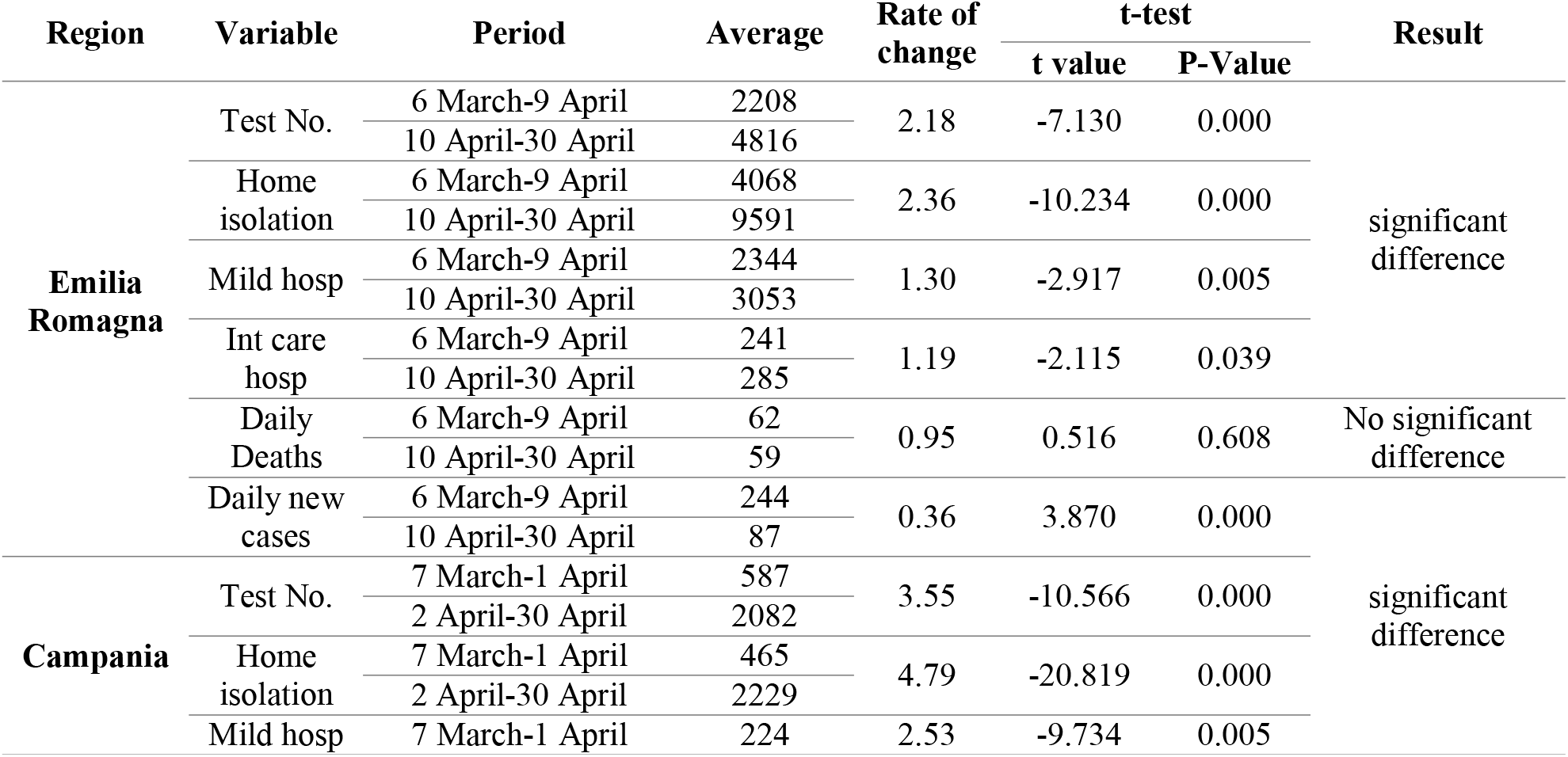
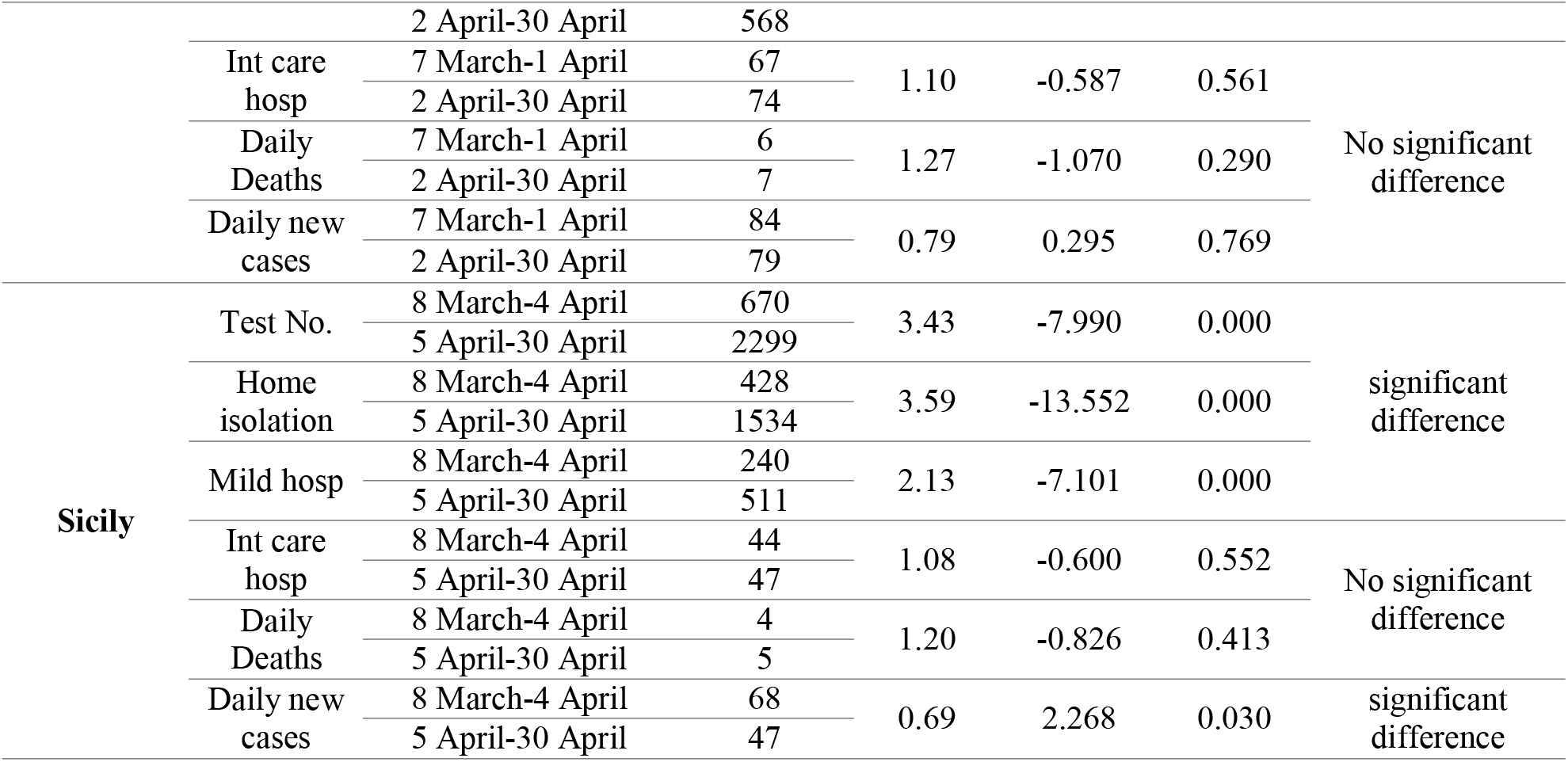
T-test results in two time periods (Emilia-Romagna, Campania, Sicily)

To summarize the T-test results of Table C3, we can say that:

- **In the three regions there has been a remarkable increase in the average number of daily tests. In Emilia-Romagna by a factor 2.18, and of the order of a factor 3.5 in Campania and Sicily, although one may observe that in the last two regions the average in the first period was quite low**.

Against this increase:

- The average number of home isolated and Mild Hosp increased significantly in all of them.
- The average number of Int Care Hosp also increased everywhere, but the increase was significant only in Emilia-Romagna.
- For what concerns Daily New Cases no significant variation was observed in Campania, differently from Emilia-Romagna and Sicily where there was a significant decrease.
- The variations in Daily Deaths were not significant

**Table C4.**
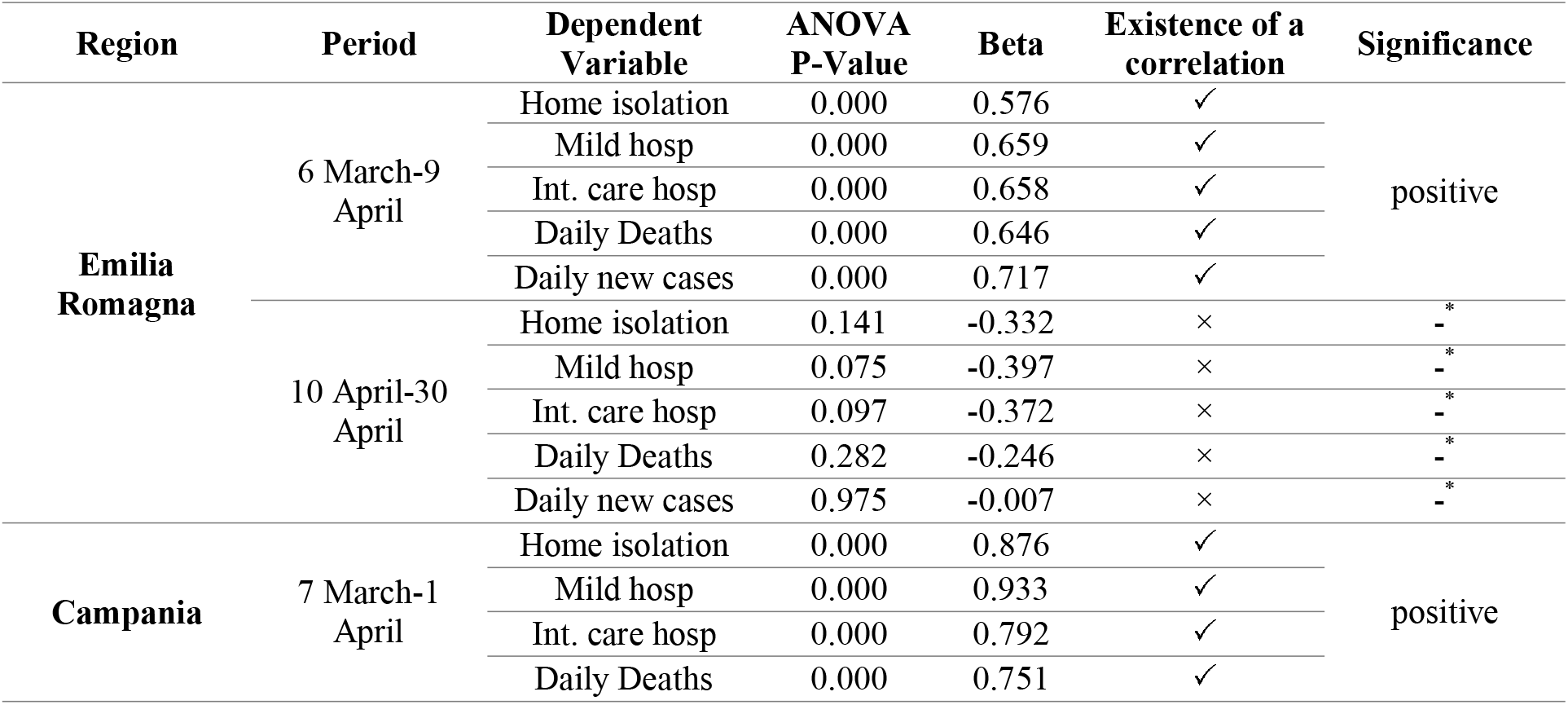
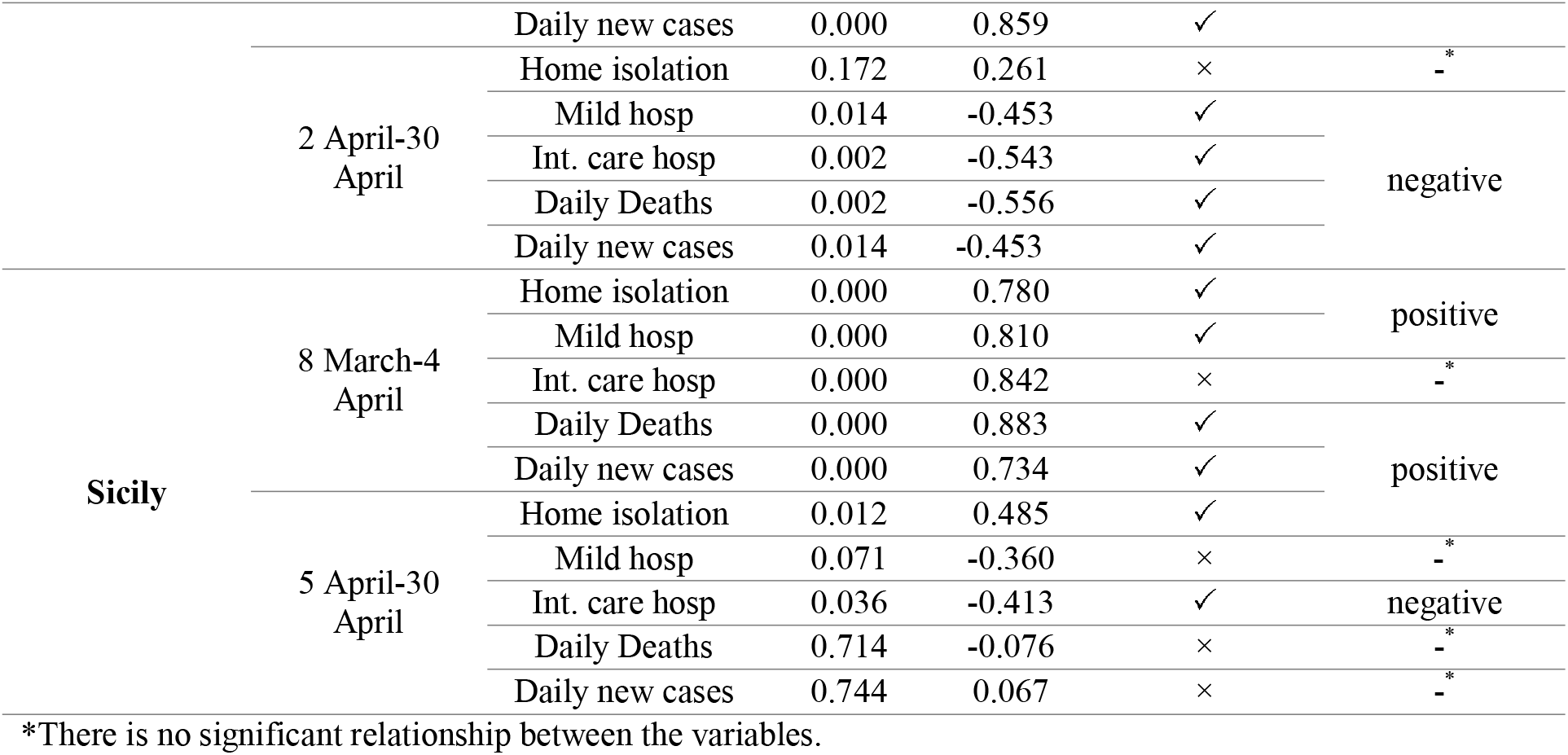
Test No. (Independent variable) Vs outcome variables in two time periods (Emilia Romagna, Campania, Sicily)

The correlation analysis of Table C4. can be summarized as follows:

- In the first period, in each region the global Italian result was confirmed with a significantly positive correlation between the number of swabs and that of each outcome variable. Sicily to some extent is the only partial exception, since the relationship for Int.care hosp, although positive, did not reach the level of being significant.
- In the second period, in the case of Emilia-Romagna there was a general turn from positive to negative relationship, but without arriving at the level of being significant. In Campania, Home isolation was positively, but not significantly correlated, whereas all the other variables exhibited a significantly negative correlation. In Sicily we found two cases of significant correlation (Home isolation, positive, and Int. care hosp, negative) and a no significant indication of negative correlation for the remaining three variables.

## Appendix D

### The critical care units in different countries

The data for other countries can be found in the studies of Prine and Wunsch (2012), and Rhodes and Moreno (2012) [57, 58]

**Table D1.** Critical care units in different countries (per 100,000 inhabitants) [56]

| Countries | Critical care units |
| --- | --- |
| United States | 34.7 |
| Germany | 29.2 |
| Italy | 12.5 |
| France | 11.6 |
| South Korea | 10.6 |
| Spain | 9.7 |
| Japan | 7.3 |
| United Kingdom | 6.6 |
| China | 3.6 |
| India | 2.3 |

